# Precision Grounding: Augmenting Large Language Models with Evidence-Based Databases for Trustworthy Genetic Variant Summarization

**DOI:** 10.1101/2025.06.09.25329279

**Authors:** Xinsong Du, Anna Nagy, Michael F. Oates, Yifei Wang, Xinyi Wang, Joseph M. Plasek, Samuel J. Aronson, Matthew S. Lebo, Li Zhou

## Abstract

**Backgrounds:** Accurate interpretation of genetic variants is critical for precision medicine. While large language models (LLMs) show promise for summarization, they are prone to hallucinations. In this study, we thus propose a novel approach named “precision grounding” that augments LLMs with a query tool that integrated evidence-based, variant-specific information to improve summarization accuracy.

**Methods:** Unlike traditional RAG methods that retrieve information via document embeddings from a vector database, precision grounding uses a domain-specific query tool to access evidence-based databases with unique identifiers. For variant summarization, we developed CATT (https://shorturl.at/pw81X), an open-source tool integrating ClinGen, ClinVar, and GenCC data. Users can query and retrieve curated evidence via Variation IDs to ground LLM outputs. We compared our approach to web grounding-based RAG using 50 expert-selected variants.

**Results:** GPT-4o was selected due to its good performance on our task during a pilot test. Using GPT-4o, we found our precision grounding approach outperformed web-search grounding, achieving significantly higher accuracy and completeness scores, which were based on a 5-point Likert-Scale of 4.76 (+0.74) and 4.94 (+0.84), respectively. Error analysis revealed that precision grounding reduced clinically significant hallucinations, such as incorrect pathogenicity classification and summarizing the wrong variant.

**Conclusion:** Precision grounding approach outperformed web-search grounding for genetic variant summarization. Our open-source tool, CATT, enables integration of curated, domain-specific knowledge and reduces hallucinations in LLM outputs.

## 1. Introduction

Genomic variation plays a significant role in human disease, with approximately 5 million variants present in each individual’s genome.^1–3^ Technological advances have made it routine to detect these variants, fueling progress in both basic and clinical research.^4^ However, the ability to detect DNA variation has outpaced our capacity to interpret the clinical relevance of these variants.^5^ Current practices focus on only a small subset of variants, limiting the potential impact of genetic testing and its contribution to research.^1^ This gap is partly due to the high personnel costs required for the in-depth assessment and summarization of evidence related to specific genes and variants.^6^ Fortunately, large language models (LLMs) hold significant potential to enhance clinical practice by automating the interpretation and summarization of genetic variants, thereby reducing costs and improving accessibility to genetic insights.

However, LLMs are prone to generating inaccurate or misleading information—a phenomenon known as hallucination.^7,8^ While many retrieval-augmented generation (RAG) techniques have been proposed to reduce hallucinations by incorporating external factual information such as Wikipedia,^9^ generic web grounding based forms of RAG retrieve information from the internet using the same techniques and internet resources across all domain and tasks.^10–13^ This grounding approach may fail to retrieve specialized, domain-specific knowledge accurately, limiting its ability to ensure factual accuracy, especially in high-stakes domain.^12,13^ In genomic medicine, such hallucinations will limit the ways these tools can be used, as incorrect variant interpretations may lead to incorrect pathogenicity classifications, flawed clinical summaries, and inconsistent evidence representation. These errors can have serious consequences in clinical decision-making, potentially leading to unnecessary medical interventions, misdiagnoses, or missed opportunities for timely treatment.

To address these challenges, we propose a “precision grounding” approach that enhances LLM with publicly available evidence-based databases and resources identified by domain experts. Unlike traditional web grounding-based RAG which uses a vector database for querying, precision grounding leverages a query tool which allows users to query relevant information based on identifiers in domain-specific databases (e.g., genetic variation ID). In the context of generic variant summarization, we developed CATT (ClinGen AI data Transformation Tool) as the query tool. Notably, CATT harmonized ClinGen,^1^ ClinVar,^8^ and GenCC^9^ as resources for retrieval augmentation and allows users to query factual information using Variation IDs in ClinVar. By implementing precision grounding, we aim to improve the factual accuracy, consistency, and clinical relevance of LLM-generated interpretations. This work fills a critical gap in applying LLMs to genomic medicine and supports safer, more reliable, and evidence-informed use of generative models in clinical decision support.

## 2. Methods

### 2.1.CATT

We developed CATT to integrate and harmonize expert curated genetic databases, including ClinGen, ClinVar, and GenCC, which are developed to support the understanding of relationships between human genetic variation and health. CATT is open-source and publicly accessible via the ClinGen website (https://shorturl.at/pw81X) and GitHub (https://github.com/mgbpm/clingen-ai-tools). Data from these sources are linked using standardized identifiers such as HGNC gene IDs or ClinVar Variation IDs to ensure consistency across datasets. Users can easily query the integrated database and retrieve information for specified variants by using command-line and specifying variation IDs. Then, CATT outputs queried genetic and clinical data into formats optimized for LLMs and other AI algorithms. The tool generates primarily two types of output: template-based natural language text and structured CSV files, both designed for direct use in LLM and AI workflows. This dual-output design enables flexible incorporation of evidence-based knowledge into prompt engineering or downstream AI applications potentially in the future enabling more robust variant interpretation and/or clinical decision support. Notably, CATT’s modular architecture supports future extensibility; new data sources can be incorporated by adding a subdirectory to the “sources” folder, with metadata and download specifications provided through a standardized set of files (“config.yaml”, “dictionary.csv”, and “mapping.csv”). Additionally, CATT supports batch processing, allowing querying multiple variants in a single run.

#### 2.1.1. Template-Based Text Generation

The tool converts structured genetic and clinical data into natural language descriptions using Genshi Template (https://genshi.readthedocs.io/en/latest/upgrade.html#text-templates), which is a flexible, standardized, and well-maintained tool. This approach allows for the dynamic generation of text summaries that preserve critical contextual and clinical details. Each dataset field, such as gene name, variant classification, or clinical significance, is mapped to a corresponding narrative structure. Users can define custom templates to specify how these structured data points should be expressed in natural language.

During processing, the tool applies the predefined templates to individual records, generating text outputs. The templates incorporate conditional phrasing, allowing the generated text to adapt based on field values. This provides context for the content, particularly for categorical and numerical data. The final text representations can be saved as separate text files for each record or concatenated into a single document.

#### 2.1.2. CSV-Based Data Representation

In addition to generating templated-based text files, CATT provides an option to structure genetic variant data into CSV format. This structured format allows LLMs to analyze tabular genetic information in conjunction with natural language queries, particularly in attachment-based input workflows where LLMs process structured data as supplementary input.

The tool allows users to specify which gene- and variant-level features should be included in the CSV output, such as gene symbol, variant ID, clinical significance, and pathogenicity classification. The column formatting is standardized to ensure consistency across records, making the data easily interpretable by LLMs. Categorical data can be converted into human-readable text to facilitate structured reasoning without requiring additional preprocessing. The CSV output can be used directly as an attachment for LLM queries, enabling models to extract insights from structured clinical and genomic records.

### 2.2. Precision Grounding to Augment LLMs to Increase Variant Summarization Trustworthiness

As illustrated in **Figure 1**, unlike the generic RAG method which first creates a vector database and then retrieves relevant information based on the similarity of document embeddings, our method first integrates expert-selected evidence-based resources then retrieves relevant information based on identifiers in domain-specific databases. Unlike generic RAG method where a vector database is needed, our precision grounding approach requires the creation of a domain-specific query tool that allows users to query information from integrated domain-specific databases. In our case (**Figure 2**), which is the genetic variant summarization task, CATT is the query tool. Our pipeline includes three steps: Step 1: Query CATT for user specified genetic variant information and get retrieved factual information in .txt and .csv files. Example .txt and .csv files are available in our GitHub repository (https://shorturl.at/tSQvK). Step 2: Upload these files as attachments into the LLM interface. Step 3: Prompt the LLM to generate a clinically useful variant summary.

**Figure 1.**
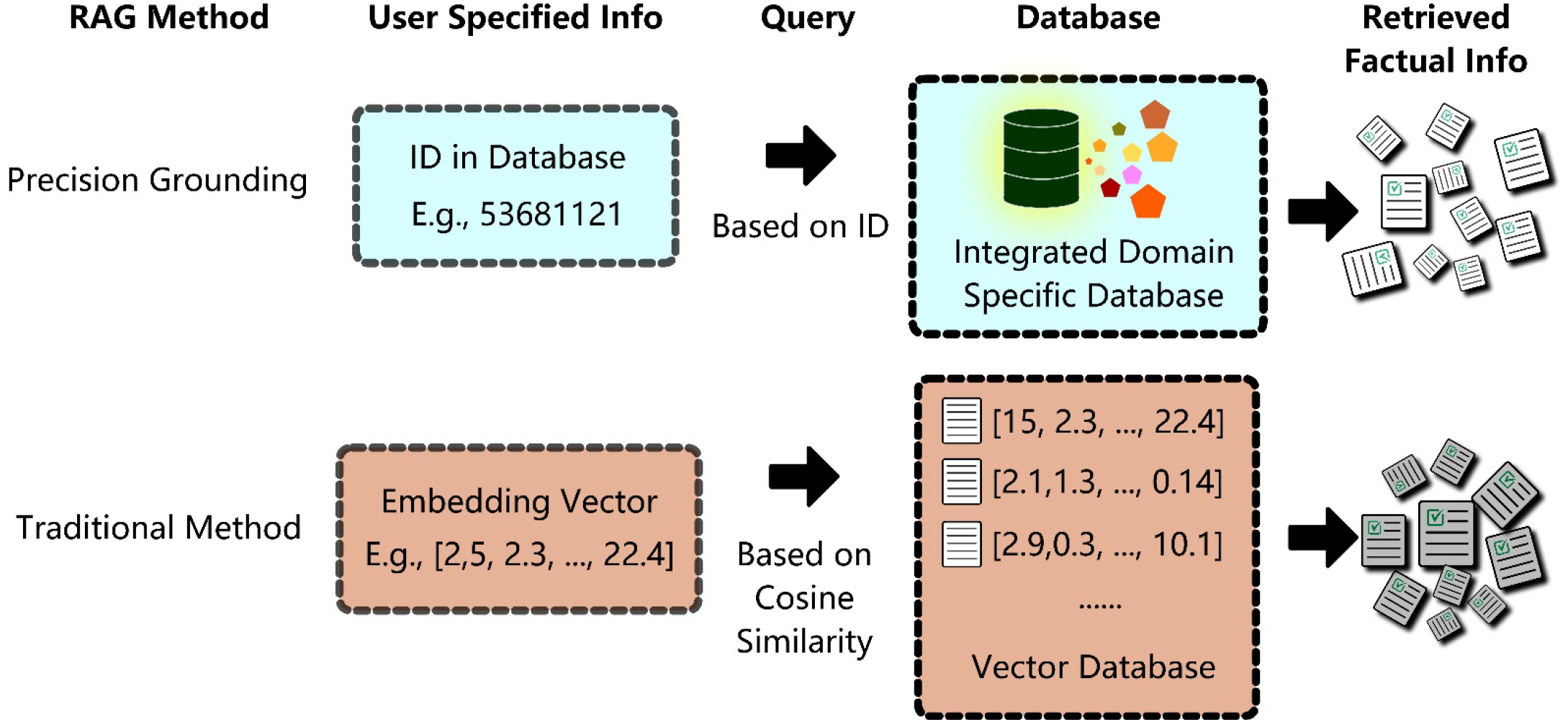
RAG Methods Description. Unlike traditional RAG methods which usually create a vector database and retrieve factual information based on cosine similarity, our proposed precision grounding method creates an integrated domain specific database, and retrieves relevant factual information based on identifiers in databases.

**Figure 2.**
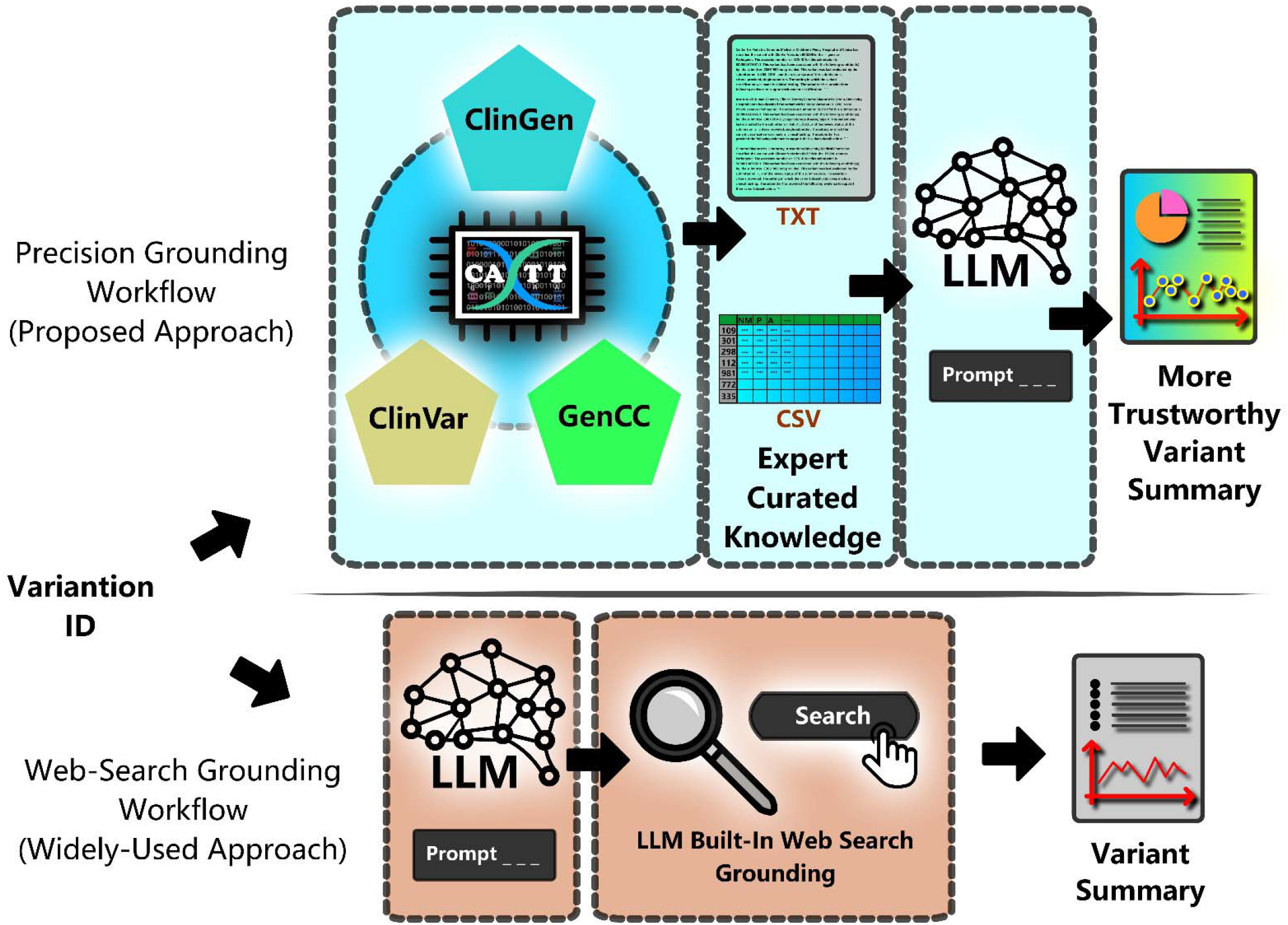
Precision Grounding vs Web-Search Grounding for Augmenting LLMs in Genetic Variant Summarization. In the traditional method, users directly prompt the LLM with web-search functionality enabled. Our precision grounding approach involves three steps: (1) generating curated variant-specific information in .txt and .csv formats using the CATT tool, (2) attaching these files to the LLM as background context, and (3) prompting the LLM to summarize the variant based on the grounded evidence.

### 2.3. Evaluation of The Effectiveness of Precision Grounding

The evaluation consisted of two phases: 1) LLM selection, 2) evaluation method selection, and 3) comparison and evaluation of precision grounding and web-search grounding. During LLM selection, we aim to select the most accurate LLM when precision grounding was not employed. Additionally, given there are a variety of evaluation approaches for assessing LLM performances,^14^ we tested and selected the most appropriate evaluation method. In the last phase, we assessed the performance of precision grounding using the selected LLM and evaluation method. We also evaluated our baseline method: web-search grounding, which has been integrated into many LLMs as a function and widely used in daily life. We compared precision grounding with web-search grounding.

A total of 50 representative genetic variants were selected for the purpose of testing and evaluation, with 10 variants used in the first and second phases and all 50 variants used in the third phase. Variant selection was designed to capture the diversity of existing variant spectrum, considering factors such as variant type (e.g., loss of function, missense, splicing), ClinVar pathogenicity classification (e.g., benign, pathogenic, risk allele), submitter concordance or discordance in ClinVar, and information density (measured by the number of gene and variant classification submissions).

#### 2.3.1. LLM Selection

We first identified the most accurate LLM for genetic variant summarization by evaluating four widely used LLMs, including GPT-4o, O1, Claude 3.5 Sonnet, and DeepSeek R1. As illustrated in **Supplementary Figure S1**, one-shot prompting was employed, as preliminary observations indicated that zero-shot prompting often produced lengthy and clinically less useful outputs. Each LLM was prompted to generate a summary intended to assist clinicians in making informed clinical decisions. The exemplary summary included in the prompt was crafted by a genetic expert to guide the LLMs toward producing clinically useful outputs.

#### 2.3.2. Evaluation Method Selection

##### 2.3.2.1. Human Evaluation

The evaluation guideline was developed through an expert consensus between two genetic experts (AN and MSL). To minimize subjective bias, LLMs’ responses were anonymized and randomized during evaluation, ensuring the reviewers were blinded to the identity of the model that generated each response.

Accuracy was manually evaluated using a 5-point Likert-Scale, and a higher accuracy means less clinically harmful.^15^ A score of 1 indicated very poor accuracy, characterized by major errors that could lead to harmful interpretation of the variant’s information. A score of 2 reflected poor accuracy with notable errors impacting understanding. A score of 3 corresponded to moderate accuracy, with inaccuracies primarily affecting supporting details rather than key findings. A score of 4 indicated minor inaccuracies or hallucinations that did not significantly alter interpretation. A score of 5 denoted perfect accuracy, devoid of any errors.

Completeness was similarly assessed using a 5-point Likert-Scale.^15^ A score of 1 indicated a complete absence of relevant information from key sources (ClinVar, ClinGen, or GenCC). A score of 2 reflected significant omissions that impaired the interpretation of the variant and/or associated gene. A score of 3 indicated partial completeness, with information drawn from at least one of the sources but lacking key elements. A score of 4 denoted mostly complete responses, with only minor omissions. A score of 5 indicated full completeness, incorporating all relevant variant and gene information from ClinVar, ClinGen, and GenCC, including American College of Medical Genetics and Genomics (ACMG) codes,^16^ pathogenicity, and gene- or disease-level details.

##### 2.3.2.2. Automated NLP Evaluation

Although human evaluation is considered the gold standard for assessing LLM performance on clinical tasks,^17,18^ it is costly, labor intensive, and time-consuming (i.e., not scalable). Therefore, we assessed the effectiveness of traditional NLP metrics in evaluating the LLM-generated summaries for genetic variants. We calculated the correlation between automated NLP metric scores and human evaluation scores using Spearman’s rank correlation coefficient, which was selected due to its widespread application in previous studies examining the relationship between human assessments and automated NLP metrics.^17,19^

#### 2.3.3. Comparison and Evaluation of Precision Grounding and Web-Search Grounding

Following model selection, we augmented the best performing model with precision grounding and web-search grounding separately. During precision grounding, the CATT-generated .CSV and .TXT files were uploaded as attachments, and the model was informed of their presence via the additional instruction section of the prompt. Regarding web-search grounding, we turned on the online-search function when prompting the LLM. Using the selected evaluation method, we then assessed the accuracy and completeness of the LLM-generated variant summaries of the two RAG methods.

### 2.4. Error Analysis

During error analysis, we categorized errors into two types based on their potential impact on clinical interpretation: 1) Clinically significant errors: these errors could alter the variant classification in a way that may lead to inappropriate clinical decisions. This category includes factual inaccuracies or hallucinations that misrepresent the variant’s pathogenicity when compared to the current knowledge (e.g., from ClinVar). For example, if the LLM incorrectly states that a benign variant is pathogenic, this could result in unnecessary interventions or anxiety for patients. 2) Clinically negligible errors: these are minor inaccuracies or misstatements that do not affect the overall interpretation or classification of the variant. Examples of these errors include numerical errors or suggesting a variant classification code that is inappropriate while maintaining the correct overall pathogenicity. Such errors are unlikely to mislead a clinician or influence medical decision-making, as the core pathogenicity assessment remain aligned with current evidence.

## 3. Results

### 3.1. Evaluation of The Effectiveness of Precision Grounding

Characteristics of the 50 included genetic variants are illustrated in **Supplementary Table S1**, which includes their pathogenicity information, and their information density in databases.

#### 3.1.1. LLM Selection

The evaluation result is illustrated in **Supplementary Table S2**. GPT-4o demonstrated the highest average accuracy (4.7), followed by DeepSeek R1 (4.1), OpenAI o1 (3.5), and Claude 3.5 Sonnet (3.5). For completeness, DeepSeek R1 scored the highest (4.9), followed by GPT-4o (4), o1 (3.8), and Claude 3.5 Sonnet (3.2). Since GPT-4o achieved the highest accuracy, it was selected to test the effectiveness of CATT. Detailed prompt and responses from LLMs during this process are illustrated in **Supplementary Table S3. Supplementary Table S4** contains notes and scores of each variant from our genetic expert when evaluating the LLM’s accuracy. Gold standard genetic summaries written by our genetic expert (AN) are provided in the **Supplementary Table S5**.

#### 3.1.2. Evaluation Method Selection

We found that all automated NLP metrics exhibited low Spearman correlations with human evaluations (**Figure 3**). For accuracy, the highest (though weak) positive correlation was observed with ROUGE-2 (ρ = 0.1295, p = 0.3702), followed by AlignScore-base^20^ (ρ = 0.1490, p = 0.3018); however, neither correlation was statistically significant, indicating a weak and unreliable association with human-rated accuracy. In contrast, for completeness, several metrics showed statistically significant negative correlations with human ratings: BLEU-2 (ρ = –0.3435, p = 0.0146), BLEU-4 (ρ = –0.3321, p = 0.0185), METEOR (ρ = –0.3134, p = 0.0267), and ROUGE-L (ρ = –0.3087, p = 0.0292). These findings indicate that none of the automated metrics reliably captured human judgements of summary accuracy. Moreover, the significant inverse correlations with completeness suggest that, in the genetic domain, lower BLEU, METEOR, and ROUGE-L scores may correspond to higher human-perceived completeness—possibly reflecting limitations of these metrics in evaluating abstractive, semantically rich summaries. Therefore, due to the weak correlation between NLP metrics with gold standard human evaluation, we decided to use human evaluation.

**Figure 3.**
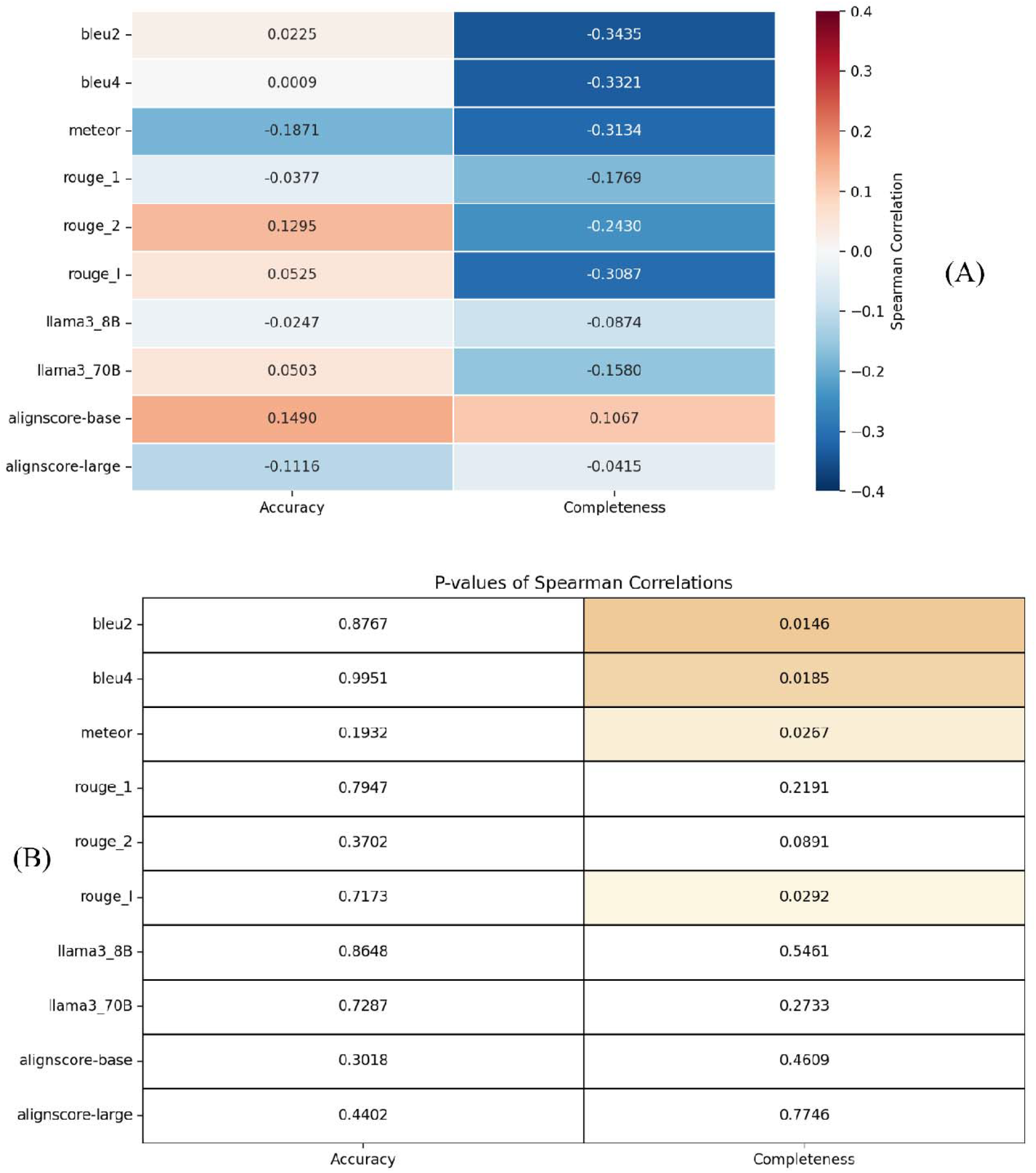
Correlation between Human Evaluation and Automated NLP Evaluation. (A) shows the Spearman correlation coefficients between human ratings (accuracy and completeness) and automated NLP metrics; (B) presents the corresponding p-values indicating statistical significance. Overall, the correlations with human-judged accuracy were weak and non-significant across all metrics (e.g., BLEU-2: ρ = 0.0225, p = 0.8767; AlignScore-base: ρ = 0.1490, p = 0.3018). In contrast, several metrics exhibited statistically significant significant correlations with completeness, though all were negative: BLEU-2 (ρ = –0.3435, p = 0.0146), BLEU-4 (ρ = –0.3321, p = 0.0185), METEOR (ρ = –0.3134, p = 0.0267), and ROUGE-L (ρ = –0.3087, p = 0.0292).

#### 3.1.3. Comparison and Evaluation of Precision Grounding and Web-Search Grounding

**Table 1** shows the performance of the best-performing model (GPT 4o) on 50 variants with precision grounding and web-search grounding. We found precision grounding could largely improve the accuracy from 4.02 to 4.76 and completeness from 4.10 to 4.94. In addition, precision grounding eliminated clinically harmful hallucination such as summarization of the wrong variant or misinterpretation of the pathogenicity of a variant. Detailed prompt and responses from LLMs during this process are illustrated in **Supplementary Table S6. Supplementary Table S7** contains notes and scores of each variant from our genetic expert during the evaluation.

**Table 1.** Manual Evaluation of LLM Performance on Genetic Variant Summarization During Model Selection (n =10 variants)

| Evaluation | GPT-4o + Web Search Grounding | o1 | Claude 3.5 Sonnet | DeepSeek R1 |
| --- | --- | --- | --- | --- |
| Average accuracy | 4.7 | 3.5 | 3.5 | 4.1 |
| Average completeness | 4.1 | 4.0 | 3.3 | 4.8 |

### 3.2 Error Analysis

Based on the evaluation of the ten variants during the LLM selection phase, as illustrated in **Supplementary Table S8**, we found O1 could misclassify the genetic variant pathogenicity and miss information from ClinVar when summarizing genetic variant. Claude 3.5 Sonnect could classify the pathogenicity incorrectly, report a wrong number of entries, and miss information from ClinVar. DeepSeek R1could also incorrectly classify the pathogenicity and report wrong number of entries. The most accurate model GPT-4o could misclassify the pathogenicity as well. Additionally, GPT 4o’s summarization sometimes misses disease information, information from ClinVar, or minor interpretation information such as ACMG code. In addition to aforementioned errors and hallucinations, during the phase of testing the effectiveness of precision grounding using 50 variants, we found both GPT-4o + web-search grounding and GPT-4o + precision grounding could overstate the pathogenicity. Nevertheless, GPT-4o + web-search grounding sometimes summarized the wrong variant (**Table 2**). Precision grounding reduced clinically harmful hallucination, including pathogenicity misclassification and summarizing the wrong variant. Additionally, precision grounding made the summary much more completed, which further improved its trustworthiness and clinical utility.

**Table 2.** Error Analysis.

| Measures | Error Category | Specific Error | Example Summary |  |
| --- | --- | --- | --- | --- |
|  |  |  | GPT-4o +<br>Web-Search<br>Grounding | GPT-4o +<br>Precision<br>Grounding |
| Accuracy | Clinically<br>significant errors | Incorrect classification of pathogenicity | X |  |
|  |  | Summarized wrong variant | X |  |
|  | Clinically<br>negligible errors | Overstated pathogenicity | X | X |
|  |  | Incorrect population frequency | X | X |
|  |  | Incorrect codes | X | X |
|  |  | Report incorrect number of submissions in databases | X |  |
| Completeness |  | Missing disease information | X |  |
|  |  | Missing information of entries in a database | X |  |
|  |  | Missing ID information (e.g., ACMG code) | X |  |
| “X” means that the model made the specific type of error(s) |  |  |  |  |

## 4. Discussion

LLMs have demonstrated remarkable capabilities in natural language understanding and generation; however, they are prone to producing content that appears factual but is ungrounded—a phenomenon known as hallucination. In genomic medicine, such inaccuracies can lead to misinterpretations of genetic variants, potentially resulting in erroneous clinical decisions. To reduce this risk, various RAG techniques have been proposed for enhance LLM outputs by incorporating external information.^9^ Nevertheless, generic web grounding-based forms of RAG retrieve information from the internet using the same techniques across all domains, limiting domain relevance and the effectiveness of hallucination reduction. In this study, we proposed precision grounding approach, which uses a query tool to retrieve context-specific evidence from expert identified databases to guide LLM outputs. We developed CATT, an open-source tool that integrates authoritative variant databases and enables users to query and retrieve relevant information for specific variants using Variation IDs. When applied to variant summarization tasks, CATT-based precision grounding significantly improves both accuracy and completeness while reducing clinically harmful hallucinations. This improvement facilitates the clinical utility of precision grounding and LLM, offering an important step toward providing healthcare professionals more trustworthy and actionable variant interpretations.

Precision grounding substantially improves the accuracy and completeness of LLM outputs for genetic variant summarization. Among the evaluated models, GPT-4o achieved the highest accuracy. This is partially attributed to its ability to perform live web searches, unlike O1, Claude 3.5, and DeepSeek R1, which did not have the function of web-search grounding at the time we used them. Notably, O1, Claude 3.5 Sonnet, and DeepSeek R1 occasionally misclassified pathogenicity of a variant, which could lead to harmful consequences in clinical practice, underscoring the need for factual grounding in LLM outputs. Despite GPT-4o’s superior performance, hallucination persisted, including misclassifying pathogenicity and summarizing the wrong variant. However, when we replaced web-based retrieval with CATT-based precision grounding, these hallucinations were eliminated. Regarding completeness, our precision grounding approach provides an LLM with comprehensive domain specific data from curated databases. This enables the LLM to receive critical annotations such as ACMG codes and disease association information, which were frequently missed by using web-search grounding. These results demonstrated the effectiveness of our precision grounding strategy and CATT toolkit in improving the reliability and comprehensiveness of LLM-generated summaries in the genetic domain.

The low correlation observed between automated NLP evaluation metrics and human assessments highlights a key limitation in current approaches to evaluating the quality of AI generated clinical text. NLP metrics, such as BLEU,^21^ ROUGE,^22^ and METEOR,^23^ primarily focus on surface-level similarity between generated and reference texts, under the assumption that good-quality outputs will have substantial lexical or structural overlap with human-written references. However, these metrics often fail to account for semantically correct paraphrasing, factual correctness, and clinical relevance – criteria that are critical to human evaluators. While human evaluation provides richer and more context-sensitive judgements, it is time-consuming, labor-intensive and subject to inter-rater variability, limiting its scalability.^24^ These challenges underscore the urgent need for more sophisticated automated evaluation methods that better align with human judgment in assessing clinical utility. Such methods would enable more efficient, consistent, and scalable evaluations of NLP models, facilitating their integration into clinical workflows where accuracy and reliability are paramount.

This study has several limitations. Due to the high cost of expert evaluation, our analysis was limited to a sample of 50 variants. Broader validation of CATT using a larger and more diverse set of variants will be necessary to comprehensively assess its effectiveness. The current version of the pipeline is not fully automated and does not include a graphical user interface, which may limit its broader adoption in clinical settings. In addition, while ClinVar and GenCC represent some of the most comprehensive publicly available resources for genetic interpretations, they are not fully curated. As a result, they may contain inaccuracies and conflicting submissions from different contributors. A comprehensive risk analysis would be required to assess the appropriateness of this tool for any clinical use. This risk analysis would have to consider current performance, bias, privacy, non-determinism as well as drift hazards, and validation is required for each specific workflow.

Future work should focus on extending precision grounding to additional domains by developing domain-specific query tools integrating expert-identified databases tailored to each domain. To improve usability and scalability, we plan to develop a fully automated pipeline that eliminates manual steps—such as extracting files from CATT and manually attaching them to LLMs via web interfaces—and enhances the software’s reproducibility and FAIRness (Findability, Accessibility, Interoperability, and Reusability).^25,26^ We also aim to implement a graphical user interface to improve CATT’s accessibility for non-technical users. The development of an automated, NLP-based evaluation framework is essential for scalable assessment of genetic variant summarization. Such a system would enable large-scale evaluation of precision grounding in the genomic domain. Additionally, methods to automatically detect hallucinations in LLM-generated variant summaries are warranted.

## 5. Conclusion

Accurate genetic variant summarization is essential for timely and reliable clinical decision-making. Although LLMs show promise for this task, hallucinations limit their clinical applicability. We proposed precision grounding and developed CATT, an open-source tool that retrieves relevant information from evidence-based databases. Our results show that the CATT-assisted precision grounding approach significantly reduces LLM hallucinations and outperforms generic web grounding-based RAG. Weak correlations between NLP metrics and expert ratings underscore the need for improved automated evaluation metrics tailored to summarization tasks. Our work is reproducible and transparent with detailed intermediate results released in supplementary materials. Though our evaluation focuses on genomics, the proposed framework is broadly extensible to other clinical domains where reliable, grounded LLM output is essential.

## Supporting information

Main Supplementary Material

## Data Availability

All data produced in the present work are contained in the manuscript

## Author Contributions

Conceptualization and design: XD, AN, MFO, SJA, MSL, LZ. Programming and software development: XD, MFO. Prompt design for large language models: XD, AN, XW, LZ. Large language model generated response preparation: XD, XW. Gold standard variant summary preparation: AN. Genetic variant summary evaluation guideline preparation and manual evaluation: AN, MSL. Calculation of correlation between natural language processing metrics and expert evaluation results: YW. Results interpretation: XD, AN, MFO, JMP, YW, SJA, LZ. Original draft preparation: XD, AN. Significant contribution to paper revision: XD, AN, SJA, LZ. Funding Acquisition: XD, MSL, LZ. Supervision: LZ.

## Conflict of Interest

None.

## Acknowledgement

Research was supported by NIH-NHGRI (U24HG006834-10S1), NIH-NIA (R01AG080429), NIH-NLM (R01LM014239), and OpenAI Researcher Access Program.

## Notes

### Competing Interest Statement

The authors have declared no competing interest.

### Summary of Updates

Revise the order of description in methods and results, making the sub-sections of the two sections more consistent

## References

1. Harrison SM, Biesecker LG, Rehm HL. Overview of Specifications to the ACMG/AMP Variant Interpretation Guidelines. Curr Protoc Hum Genet. 2019;103(1):e93. doi:10.1002/cphg.93

2. Green RC, Shah N, Genetti CA, et al. Actionability of unanticipated monogenic disease risks in newborn genomic screening: Findings from the BabySeq Project. Am J Hum Genet. 2023;110(7):1034–1045. doi:10.1016/j.ajhg.2023.05.007

3. Machini K, Ceyhan-Birsoy O, Azzariti DR, et al. Analyzing and Reanalyzing the Genome: Findings from the MedSeq Project. Am J Hum Genet. 2019;105(1):177–188. doi:10.1016/j.ajhg.2019.05.017

4. Diao JA, Kohane IS, Manrai AK. Biomedical informatics and machine learning for clinical genomics. Hum Mol Genet. 2018;27(R1):R29–R34. doi:10.1093/hmg/ddy088

5. De La Vega FM, Chowdhury S, Moore B, et al. Artificial intelligence enables comprehensive genome interpretation and nomination of candidate diagnoses for rare genetic diseases. Genome Med. 2021;13(1):153. doi:10.1186/s13073-021-00965-0

6. McInnes G, Sharo AG, Koleske ML, et al. Opportunities and challenges for the computational interpretation of rare variation in clinically important genes. Am J Hum Genet. 2021;108(4):535–548. doi:10.1016/j.ajhg.2021.03.003

7. Farquhar S, Kossen J, Kuhn L, Gal Y. Detecting hallucinations in large language models using semantic entropy. Nature. 2024;630(8017):625–630. doi:10.1038/s41586-024-07421-0

8. Du X, Novoa-Laurentiev J, Plasek JM, et al. Enhancing early detection of cognitive decline in the elderly: a comparative study utilizing large language models in clinical notes. EBioMedicine. 2024;109:105401. doi:10.1016/j.ebiom.2024.105401

9. Lewis P, Perez E, Piktus A, et al. Retrieval-augmented generation for knowledge-intensive NLP tasks. In: Proceedings of the 34th International Conference on Neural Information Processing Systems. NIPS ‘20. Curran Associates Inc.; 2020:9459–9474.

10. Masanneck L, Meuth SG, Pawlitzki M. Evaluating base and retrieval augmented LLMs with document or online support for evidence based neurology. npj Digit Med. 2025;8(1):1–5. doi:10.1038/s41746-025-01536-y

11. Wang D, Zhang S. Large language models in medical and healthcare fields: applications, advances, and challenges. Artif Intell Rev. 2024;57(11):299. doi:10.1007/s10462-024-10921-0

12. Li X, Zhao R, Chia YK, et al. Chain-of-Knowledge: Grounding Large Language Models via Dynamic Knowledge Adapting over Heterogeneous Sources. Published online February 21, 2024. doi:10.48550/arXiv.2305.13269

13. Kenthapadi K, Sameki M, Taly A. Grounding and Evaluation for Large Language Models: Practical Challenges and Lessons Learned (Survey). In: Proceedings of the 30th ACM SIGKDD Conference on Knowledge Discovery and Data Mining. KDD ‘24. Association for Computing Machinery; 2024:6523–6533. doi:10.1145/3637528.3671467

14. Du X, Zhou Z, Wang Y, et al. Adapting Generative Large Language Models for Electronic Health Record Applications: A Systematic Review of Methodologies, Evaluation, and Hallucinations. Published online May 10, 2025:2024.08.11.24311828. doi:10.1101/2024.08.11.24311828

15. Albaum G. The Likert Scale Revisited. Market Research Society Journal. 1997;39(2):1–21. doi:10.1177/147078539703900202

16. Richards S, Aziz N, Bale S, et al. Standards and guidelines for the interpretation of sequence variants: a joint consensus recommendation of the American College of Medical Genetics and Genomics and the Association for Molecular Pathology. Genet Med. 2015;17(5):405–424. doi:10.1038/gim.2015.30

17. Van Veen D, Van Uden C, Blankemeier L, et al. Adapted large language models can outperform medical experts in clinical text summarization. Nat Med. 2024;30(4):1134–1142. doi:10.1038/s41591-024-02855-5

18. Tam TYC, Sivarajkumar S, Kapoor S, et al. A framework for human evaluation of large language models in healthcare derived from literature review. npj Digit Med. 2024;7(1):1–20. doi:10.1038/s41746-024-01258-7

19. Tie X, Shin M, Pirasteh A, et al. Personalized Impression Generation for PET Reports Using Large Language Models. J Digit Imaging Inform med. 2024;37(2):471–488. doi:10.1007/s10278-024-00985-3

20. Zha Y, Yang Y, Li R, Hu Z. AlignScore: Evaluating Factual Consistency with a Unified Alignment Function. Published online May 26, 2023. doi:10.48550/arXiv.2305.16739

21. Papineni K, Roukos S, Ward T, Zhu WJ. Bleu: a Method for Automatic Evaluation of Machine Translation. In: Isabelle P, Charniak E, Lin D, eds. Proceedings of the 40th Annual Meeting of the Association for Computational Linguistics. Association for Computational Linguistics; 2002:311–318. doi:10.3115/1073083.1073135

22. Lin CY. ROUGE: A Package for Automatic Evaluation of Summaries. In: Text Summarization Branches Out. Association for Computational Linguistics; 2004:74–81. Accessed March 2, 2025. https://aclanthology.org/W04-1013/

23. Banerjee S, Lavie A. METEOR: An Automatic Metric for MT Evaluation with Improved Correlation with Human Judgments. In: Goldstein J, Lavie A, Lin CY, Voss C, eds. Proceedings of the ACL Workshop on Intrinsic and Extrinsic Evaluation Measures for Machine Translation and/or Summarization. Association for Computational Linguistics; 2005:65–72. Accessed March 2, 2025. https://aclanthology.org/W05-0909/

24. Croxford E, Gao Y, Pellegrino N, et al. Current and future state of evaluation of large language models for medical summarization tasks. npj Health Syst. 2025;2(1):1–13. doi:10.1038/s44401-024-00011-2

25. Du X, Dobrowolski A, Brochhausen M, Garrett TJ, Hogan WR, Lemas DJ. Nextflow4MS-DIAL: A Reproducible Nextflow-Based Workflow for Liquid Chromatography-Mass Spectrometry Metabolomics Data Processing. J Am Soc Mass Spectrom. Published online January 5, 2025. doi:10.1021/jasms.4c00364

26. Du X, Dastmalchi F, Ye H, et al. Evaluating LC-HRMS metabolomics data processing software using FAIR principles for research software. Metabolomics. 2023;19(2):11. doi:10.1007/s11306-023-01974-3

