## Supplementary material for "Precision Grounding: Augmenting Large Language Models with Evidence-Based Databases for Trustworthy Genetic Variant Summarization": Main Supplementary Material

**Supplementary Tables**

**Table S1.** Included Genetic Variants

| Variation ID | Name | Pathogenicity Classification | ClinGen gene information available | GenCC gene information available | ClinVar variant information available |
| --- | --- | --- | --- | --- | --- |
| 457630 | NM_001042492.3(NF1):c.3198-3C>T | VUS/LB/B | Classification, dosage sensitivity and actionability information available | 5 submitters | 3 submissions, 1 with evidence provided |
| 323645 | NM_001377265.1(MAPT):c.1630G>A (p.Ala544Thr) | VUS/LB/B/Risk allele | Dosage sensitivity information available | 2 submitters | 6 submissions, 5 with evidence provided |
| 157939 | NM_000053.4(ATP7B):c.2605G>A (p.Gly869Arg) | P/LP | Classification, dosage sensitivity and actionability information available | 4 submitters | 27 submissions, 14 with evidence |
| 5760 | NM_032551.5(KISS1R):c.1157G>C (p.Arg386Pro) | LP/VUS/LB | No information | 3 submitters | 4 submission, 1 with evidence provided |
| 438635 | NM_023936.2(MRPS34):c.94C>T | P/LP | Classification information available | 2 submitters | 14 submissions, 7 with evidence provided |
| 402592 | NM_177438.3(DICER1):c.4206+9_4206+11del | LB/B | Classification, dosage sensitivity and actionability information available | 4 submitters | 7 submissions, 2 with evidence provided |
| 42454 | NM_000169.3(GLA):c.352C>T (p.Arg118Cys) | VUS/LB | Classification, dosage sensitivity and actionability information available | 4 submitters | 25 submissions, 11 with evidence provided |
| 2298 | NM_005609.4(PYGM):c.148C>T (p.Arg50Ter) | P | ClinGen actionability information available | 5 submitters | 34 submissions, 15 with evidence provided |
| 1027708 | NM_138348.6(OTULIN):c.22C>T (p.Gln8Ter) | P | No information | 2 submitters | 1 submissions, no evidence provided |
| 8431 | NM_000304.4(PMP22):c.353C>T (p.Thr118Met) | VUS/LB/B | Classification, dosage sensitivity and actionability information available | 6 submitters | 25 submissions, 6 with evidence provided |
| 13507 | NM_002693.3(POLG):c.2243G>C (p.Trp748Ser) | P/LP | Classification, pharmacogenomics | 6 submitters | 43 submissions, 16 with evidence provided |
| 228294 | NM_001039876.3(SYNE4):c.699G>A (p.Trp233Ter) | P/LP/VUS | Classification and dosage sensitivity | 4 submitters | 7 submissions, 4 with evidence provided |
| 370854 | NM_000055.4(BCHE):c.635C>T (p.Ala212Val) | P/LP/VUS | Dosage sensitivity and pharmacogenomics | 4 submitters | 7 submissions, 6 with evidence provided |
| 225370 | NM_001486.4(GCKR):c.307G>A (p.Val103Met) | LP/VUS/B | No information | Not present | 4 submissions, 1 with evidence provided |
| 195696 | NM_001384732.1(CPLANE1):c.3828T>C (p.Leu1276=) | VUS/LB/B | Classification and dosage sensitivity | 5 submitters | 9 submissions, 5 with evidence provided |
| 381621 | NM_000552.5(VWF):c.1625C>G (p.Ala542Gly) | P/LP/VUS | Classification, actionability | 4 submitters | 10 submissions, 6 with evidence provided |
| 360767 | NM_001001548.3(CD36):c.1133G>T (p.Gly378Val) | LP/VUS | Dosage sensitivity | 2 submitters | 3 submissions, 2 with evidence provided |
| 310 | NM_000552.5(VWF):c.4751A>G (p.Tyr1584Cys) | P/LP/VUS | Classification, actionability | 4 submitters | 32 submissions, 13 with evidence provided |
| 8046 | NM_005055.5(RAPSN):c.264C>A (p.Asn88Lys) | P/LP | No information | 4 submitters | 41 submissions, 16 with evidence provided |
| 222816 | NM_005138.3(SCO2):c.16_17insAGCATGCAGCAGTGACTCA (p.Arg6fs) | P/LP | Classification | 3 submitters | 7 submissions, 3 with evidence provided |
| 1285628 | NM_001204375.2(NPR3):c.442T>C (p.Ser148Pro) | P/LP | No information | 1 submitter | 2 submissions, 1 with evidence provided |
| 1429622 | NM_001243279.3(ACSF3):c.1609del (p.Ala537fs) | P/LP | Classification and dosage sensitivity | 3 submitters | 2 submissions, 1 with evidence provided |
| 632033 | NM_005502.4(ABCA1):c.1759C>T (p.Arg587Trp) | P/LP | No information | 4 submitters | 3 submissions, 3 with evidence provided |
| 92870 | NM_000350.3(ABCA4):c.5461-10T>C | P/LP | Classification | 6 submitters | 40 submissions, 14 with evidence provided |
| 191059 | NM_014249.4(NR2E3):c.119-2A>C | P/LP | No information | 4 submitters | 37 submissions, 13 with evidence provided |
| 31811 | NM_032578.4(MYPN):c.59A>G (p.Tyr20Cys) | P/LP/VUS/LB | Classification | 5 submitters | 21 submissions, 8 with evidence provided |
| 197403 | NM_213599.3(ANO5):c.155A>G (p.Asn52Ser) | LP/VUS/LB/B | Classification | 3 submitters | 12 submissions, 5 with evidence provided |
| 1205854 | NM_138289.4(ACTRT1):c.547dup (p.Met183fs) | LP/VUS/LB | No information | No information | 5 submission, 0 with evidence provided |
| 216972 | NM_000435.3(NOTCH3):c.3691C>T (p.Arg1231Cys) | P/LP/VUS/B | Classification and Actionability | 6 submitters | 16 submissions, 9 with evidence provided |
| 414302 | NM_001114753.3(ENG):c.-9G>A | LP/VUS/LB | Classification, dosage sensitivity, actionability | 6 submitters | Expert panel VUS, 14 submissions |
| 225415 | NM_017534.6(MYH2):c.2414T>C (p.Val805Ala) | LP/VUS/B | Classification | 6 submitters | 7 submissions, 4 with evidence provided |
| 41728 | NM_000548.5(TSC2):c.1458C>G (p.Asn486Lys) | VUS/LB/B | Classification, dosage sensitivity, actionability | 7 submitters | 6 submissions, 2 with evidence provided |
| 562374 | NM_004621.6(TRPC6):c.643C>T (p.Arg215Trp) | LP/VUS | No information | 3 submitters | 4 submissions, 1 with evidence provided |
| 18272 | NM_000036.3(AMPD1):c.1162C>T (p.Arg388Trp) | P/LP/VUS/LB | No information | 2 submitters | 7 submitters, 3 with evidence provided |
| 444210 | NM_006214.4(PHYH):c.1010_1012dup (p.Asn337_Leu338insHis) | LP/VUS/LB | Classification, dosage sensitivity, actionability | 5 submitters | 6 submissions, 1 with evidence provided |
| 290893 | NM_001807.6(CEL):c.2172del (p.Val725fs) | LP/VUS/B | Classification | 5 submitters | 4 submissions, 1 with evidence provided |
| 229616 | NM_206933.4(USH2A):c.12145G>A (p.Ala4049Thr) | P/LP/VUS | Classification, dosage sensitivity | 4 submitters | 10 submissions, 6 with evidence provided |
| 372641 | NM_001171.6(ABCC6):c.496C>T (p.Arg166Cys) | LP/VUS | No information | 3 submitters | 5 submissions, 2 with evidence provided |
| 459 | NM_000097.7(CPOX):c.1339C>T (p.Arg447Cys) | P/LP/VUS | Classification | 5 submitters | 6 submissions, 2 with evidence provided |
| 39974 | NM_004525.3(LRP2):c.6160G>A (p.Asp2054Asn) | P/LP/VUS/LB | Classification, dosage sensitivity | 4 submitters | 8 submissions, 4 with evidence provided |
| 50866 | NM_130837.3(OPA1):c.1311A>G (p.Ile437Met) | P/LP/VUS | Classification | 8 submitters | 20 submissions, 12 with evidence provided |
| 16451 | NM_000138.5(FBN1):c.3509G>A (p.Arg1170His) | VUS/LB/B | Classification, dosage sensitivity, actionability | 7 submitters | Expert Panel B, 34 submissions |
| 46597 | NM_001267550.2(TTN):c.1137A>G (p.Arg379=) | LP/VUS/LB/B | Classification, dosage sensitivity | 5 submitters | 22 submissions, 10 with evidence provided |
| 684532 | NM_014889.4(PITRM1):c.2647C>T (p.Leu883Phe) | B/LB | No information | 2 submitters | 5 submissions, 2 with evidence provided |
| 8678 | NM_000525.4(KCNJ11):c.67A>G (p.Lys23Glu) | B/LB | Classification | 6 submitters | 18 submissions, 6 with evidence provided |
| 17864 | NM_000041.4(APOE):c.388T>C (p.Cys130Arg) | P/LP/VUS/Risk allele | Pharmacogenomics | 4 submitters | 13 submissions, 4 with evidence submitted |
| 5603 | NM_007194.4(CHEK2):c.1283C>T (p.Ser428Phe) | P/LP/P(low pen)/Risk allele/VUS | Classification, dosage sensitivity, clinical actionability | 5 submitters | 37 submissions, |
| 575178 | NM_000059.4(BRCA2):c.6859A>T (p.Arg2287Ter) | P/LP/LP low pen | Classification, dosage sensitivity, actionability | 5 submitters | 3 submissions, 3 with evidence submitted |
| 5302 | NM_012452.3(TNFRSF13B):c.310T>C (p.Cys104Arg) | P/LP/risk factor/VUS/LB | Classification | 5 submitters | 47 submissions, 22 with evidence submitted |
| 3020178 | NM_014049.5(ACAD9):c.9C>T (p.Gly3=) | LB | Classification | 4 submitters | 1 submission, 0 with evidence submitted |
| Abbreviations: B-Benign; LB-Likely Benign; VUS-Variant of Uncertain Significance; LP-Likely Pathological; P-Pathological | | | | | |

| Table S2. Manual Evaluation of LLM (GPT-4o) Performance Using Web-Search vs. Precision Grounding (n=50 variants) | | |
| --- | --- | --- |
| Variant ID | **GPT-4o + Web Search Grounding** | **GPT-4o + Precision Grounding Using CATT** |
| Average Accuracy | 4.02 | 4.76 |
| Average Completeness | 4.10 | 4.94 |

**Table S5.** Gold Standard Summaries Written by Genetic Expert

| Variation ID | Summary |
| --- | --- |
| 562 | The NM_000140.5:c.315-48T>C variant in the FECH gene is a splice variant. The FECH gene has been definitively associated with autosomal recessive erythropoietic protoporphyria by ClinGen. In addition, the FECH gene has been associated with autosomal recessive erythropoietic protoporphyria by 5 submitters in GenCC with the majority classifying this as a strong or definitive association. In ClinVar, this variant has conflicting classifications of pathogenicity based on 18 submissions. The majority of submissions classify this variant as Likely Pathogenic or Pathogenic, however 2 submissions which were assessed in 2016 classified this as a Variant of Uncertain Significance, and 1 submission classifies this as Pathogenic with Low Penetrance. Based on submission summaries in ClinVar, this variant has been reported in the literature in individuals with erythropoietic protoporphyria. Functional studies have also demonstrated aberrant splicing and a mild reduction in enzyme activity. Multiple submissions note that this variant may not cause disease or may have lower penetrance in the homozygous state, and may only be pathogenic when found with a loss-of-function or deleterious allele. This variant has been found in population databases such as gnomAD. ACMG codes which have been applied by submitters include: PM3_VeryStrong, PP1_Strong, PS3_Moderate, PM3, PP6, BS2. |
| 157939 | The NM_000053.4:c.2605G>A (p.Gly869Arg) variant in the ATP7B gene is a missense variant. The ATP7B gene has been definitively associated with autosomal recessive Wilson disease by ClinGen. In addition, it has been associated with Wilson disease by 3 submitters in GenCC (in addition to ClinGen), with 2 submitters classifying it as Strong and Orphanet classifying it as supportive. ClinGen has also noted that there is clinical actionability for this gene in the context of Wilson Disease including interventions such as copper chelation, zinc therapy and diet. In ClinVar, this variant is classified as Pathogenic/Likely Pathogenic in ClinVar based on 25 individual submissions. Based on the submission summaries in ClinVar, this variant has been reported in the literature in individuals with Wilson disease alongside other pathogenic variants. This variant has been found in population databases such as gnomAD. In silico tools predict a damaging effect on protein function and the variant is highly conserved. Multiple submitters also note that this variant is in a well-established functional domain (ATPase domain). Multiple submitters note that this may be a mild variant with reduced penetrance or late onset disease as it has been found in asymptomatic individuals and affected. ACMG codes which have been applied by submitters include: PP3, PP4_Moderate, PM2/PM2_Supporting, PS4/PS4_Moderate, PM3_VeryStrong/PM3_Strong, PM1. |
| 323645 | The NM_001377265.1:c.1630G>A (p.Ala544Thr) variant in the MAPT gene is a missense variant. The MAPT gene has been associated with progressive supranuclear palsy, late-onset Parkinson disease, Pick disease and semantic dementia by 2 submitters in GenCC. Supranuclear palsy has an autosomal recessive inheritance pattern, and Pick disease, late-onset Parkinson disease, and semantic dementia have an autosomal dominant inheritance pattern. Pick disease has a gene-disease association of Strong by both submitters, however the other diseases have a range of associations from Limited to Strong. There is currently no evidence for haploinsufficiency or triplosensitivity by ClinGen. In Clinvar, this variant has conflicting classifications of pathogenicity from 4 individual submissions; 3 of them are Likely Benign or Benign, and 1 is Variant of Uncertain Significance (VUS). The classification of VUS is from Invitae and is the most recently evaluated assertion. The 3 submitters with Likely Benign or Benign classifications have used the BP4 and BS2 ACMG codes to classify this, however otherwise do not provide significant information about their classification. Invitae notes that this variant was reported in a single study to increased the risk for FTD, however the result has not been replicated. In addition, experimental studies have shown that the change affects protein function. At this time, there is conflicted and limited information for pathogenicity, and at most, this variant may represent a risk factor. |
| 457630 | The NM_001042492.3:c.3198-3C>T variant in the NF1 gene is a splice variant. The NF1 gene has been definitively associated with autosomal dominant neurofibromatosis type 1 by ClinGen. An additional 4 submitters have a strong or definitive association with neurofibromatosis type 1 in GenCC. Some submitters have also associated the NF1 gene with other diseases such as neurofibromatosis-Noonan syndrome, Watson syndrome, familial spinal neurofibromatosis, hereditary pheochromocytoma-paraganglioma syndrome and Moyamoya disease. There are potential interventions for this gene including surveillance for breast cancer and consultation with a provider for peripheral nerve sheath tumors. In ClinVar, this variant has conflicting classifications of pathogenicity from 3 individual submitters; 2 of them are Likely Benign/Benign and 1 submission is Uncertain Significance. No significant evidence has been provided by the submitters to support the classifications. At this time there is little evidence to support pathogenicity for this variant. |
| 1027708 | The NM_138348.6:c.22C>T (p.Gln8Ter) variant in the OTULIN gene is a nonsense variant. The OTULIN gene has been associated with autosomal recessive infantile-onset periodic fever-panniculitis-dermatosis syndrome by 3 submitters. This variant has been classified as Pathogenic in ClinVar by 1 submitter, however no evidence has been provided to support the classification. |
| 2298 | The NM_005609.4:c.148C>T (p.Arg50Ter) variant in the PYGM gene is a nonsense variant. The PYGM gene has been associated with autosomal recessive glycogen storage disease V by 4 submitters, with a classification of Strong by the majority of submissions. It also has limited association with autosomal dominant glycogen storage disease V by 1 submitter. This gene has a potential intervention of implementing an exercise regimen with moderate actionability by ClinGen. This variant has been classified as Pathogenic in ClinVar by 33 submitters. Based on submission summaries in ClinVar, this variant is one of the most common, well established pathogenic variants in this gene. It has been found in many individuals in literature who are homozygotes or compound heterozygotes. In addition, it has segregated with disease in multiple families. The variant is predicted to undergo nonsense mediated decay, and functional studies support that protein transcript was not found in individuals with this variant. Loss of function variants in this gene have been shown to be pathogenic. Mouse models show that the variant affects protein function. The variant has been found in reference populations such as gnomAD. ACMG codes which have been applied by submitters include: PVS1, PS3/PS3_Supporting, PS4, PM3/PM3_VeryStrong, PM2, PP1_Strong/PP1. |
| 42454 | The NM_000169.3(GLA):c.352C>T (p.Arg118Cys) variant in the GLA gene is a missense variant. The GLA gene has a definitive association with X-linked Fabry disease by Clingen. In addition, 3 other submitters have a strong or definitive association with Fabry disease, and Orphanet is supportive. Loss of function variants in this gene may be disease causing, as there is sufficient evidence for haploinsufficiency by ClinGen. There is no current evidence for triplosensitivity. ClinGen has identified enzyme replacement therapy as a possible intervention for this gene to treat multiple disease outcomes including end-stage renal disease, cerebrovascular events and cardiovascular disease. This variant has conflicting classification in ClinVar based on 23 individual submissions. The majority of these submissions classify the variant as a Variant of Uncertain Significance, with a few submissions classifying this as Likely Benign. One flagged submission classifies this variant as Likely Pathogenic. This variant has been found in multiple individuals in the literature with Fabry disease and was also found in affected and unaffected family members. This variant has also been found at a high frequency in reference populations such as gnomAD. Functional studies show a decreased in enzyme activity, however some submitters not that it may not reach the levels of classic Fabry. In addition, a pathogenic variant was found in cis with this variant in a family, supporting a benign role. Some submitters suggest reduced penetrance. ACMG codes which have been applied by submitters include: PS4_Moderate, PVS1_Moderate, PP1, PP3, BS4, BS1, PS3_Moderate, PP4, BP2, BS2. |
| 402592 | The NM_177438.3(DICER1):c.4206+9_4206+11del variant in the DICER1 gene is an intronic deletion of 3 nucleotides. The DICER1 gene has a definitive association with Autosomal dominant tumor predisposition. In addition, submitters in GenCC have reported a Limited association with global developmental delay-lung cysts-overgrowth-Wilms tumor syndrome, a definitive association with autosomal dominant multinodular goiter with or without Sertoli-Leydig cell tumors and definitive association with autosomal dominant pleuropulmonary blastoma. ClinGen has identified chest imaging and surveillance as possible interventions for DICER1-related cancers. Loss of function variants in this gene may be disease causing, as ClinGen has found sufficient evidence for haploinsufficiency. There is currently no evidence for triplosensitivity. In ClinVar, this variant has been classified as Likely Benign or Benign by 7 submissions, therefore this suggests the variant is not disease causing. No significant evidence has been provided by the submitters to support the classifications. |
| 438635 | The NM_023936.2(MRPS34):c.94C>T (p.Gln32Ter) variant in the MRPS34 gene is a nonsense variant. The MRPS34 gene has a moderate association to autosomal recessive Leigh syndrome by ClinGen. In addition, Invitae has found a strong association to autosomal recessive oxidative phosphorylation deficiency 32. In ClinVar, this variant has been classified as Likely Pathogenic or Pathogenic by 11 submitters, suggesting that this variant may be disease causing. Multiple submitters note that loss of function is an established mechanism of disease for this gene. This variant has been identified in an individual in the literature with Leigh syndrome. Functional studies of this variant show an effect on the protein. This variant has been found in population databases such as gnomAD. ACMG codes which have been applied by submitters include: PVS1, PM3/PM3_Supporting, PP3, PS3 |
| 5760 | The NM_032551.5(KISS1R):c.1157G>C (p.Arg386Pro) variant in the KISS1R gene is a missense variant. Multiple submitters in GenCC have associated this gene to autosomal recessive hypogonadotropic hypogonadism with or without anosmia. In addition, one submitter has reported a limited association to autosomal recessive centra precocious puberty. In ClinVar, this variant has conflicting classifications from 3 submitters, with one lab classifying this as Likely Pathogenic and one lab classifying this as Likely Benign. OMIM also classifies this as Pathogenic. Evidence supporting these classifications are only available for the likely pathogenic classification. This submission states that has been an individual in the literature with this variant and functional studies show a gain-of-function effect. This variant is not conserved across species and is not found in the NHLBI Exome Sequencing project. Based on this information it is unclear if this variant causes disease. |

| Measures | Category | Error Categories and Examples | Large Language Models | | | | |
| --- | --- | --- | --- | --- | --- | --- | --- |
|  |  |  | **OpenAI o1** | **Claude 3.5 Sonnet** | **DeepSeek R1** | **GPT-4o + Web Search Grounding** | **GPT-4o + Precision Grounding** |
| Accuracy | Pathogenicity classification inaccuracy | Incorrect classification of pathogenicity | **X** | **X** | **X** | **X** |  |
|  | Supporting detail inaccuracy | Incorrect number of entries in database | **X** | **X** | **X** | **X** | **X** |
|  |  | Overstated pathogenicity |  | **X** | **X** |  |  |
| Completeness | | Missing disease information |  |  |  | **X** |  |
|  |  | Missing information from ClinVar | **X** | **X** |  | **X** |  |
|  |  | Missing important ID information (e.g., ACMG code) |  |  |  | **X** |  |
| “X” means that the model made the specific type of error(s) | | | | | | | |

**Table S8.** Errors during LLM Selection Phase

**Supplementary Figures**

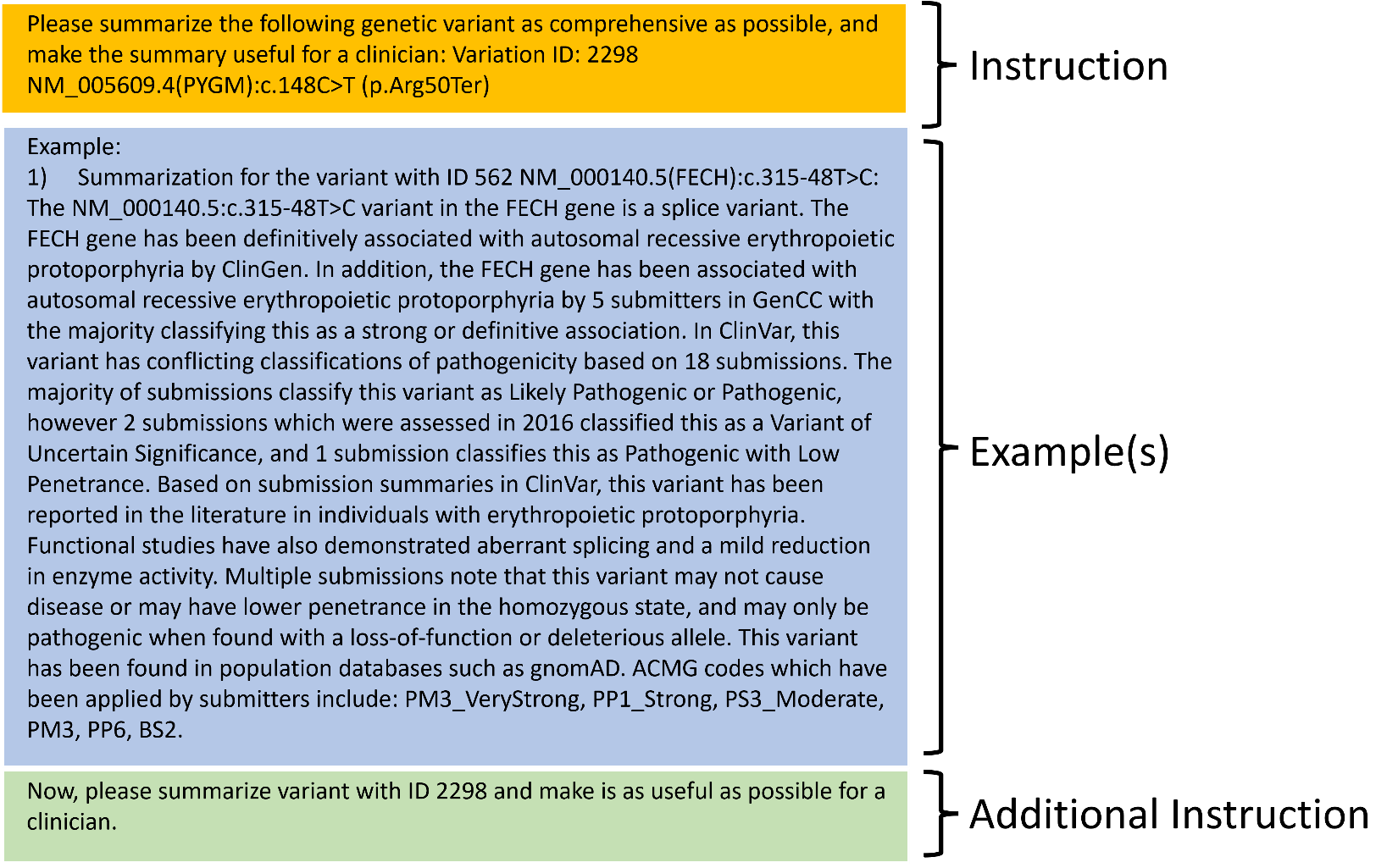

**Figure S1.** Prompt Structure. Our crafted prompt includes three sections: instruction, example(s), and additional instruction. In the instruction section, we asked the LLM to make a clinically useful summary for the specific variant, and specified the variation ID and name, since we observed that the LLM always fabricates the variant name if we only include the ID but did not include the name in the prompt. In the example(s) section, we included a gold standard summary written by our genetic expert, as we found that the LLM usually outputs a very lengthy summary with redundant information without an example. In the additional instruction section, we rephrase the task and notify the LLM to look at attachments (if there are), since we found notifying the LLM about the attachment could slightly improve the accuracy.
